# Improved AI-based Prediction of ICU Mortality of Sepsis Patients based on Change in HRV Parameter upon Fluid Bolus Therapy

**DOI:** 10.1101/2025.07.23.25332107

**Authors:** LR Rahul, Rahuldeb Sarkar, Arnab Sengupta, Soumya Jana

**Author notes:** Contributing authors.

## Abstract

**Purpose:** To examine the role of heart rate variability (HRV) changes in response to fluid bolus therapy (FBT) on mortality prediction among sepsis patients in the intensive care unit (ICU).

**Methods:** Adult sepsis patients in the Medical Information Mart for Intensive Care (MIMIC)-III and MIMIC-IV clinical database, who also had continuous ECG in the corresponding waveform database, within 48 hours of ICU admission were selected for the study. All models were developed using a common architectural template, each based on an ensemble of decision trees making use of eXtreme Gradient Boosting (XGBoost). A mortality prediction model (Model-1) was built including SAPS II score and common pool of features (CPOF) comprising of age, sex, ethnicity, insurance, admission type, heart rate, mean arterial pressure (MAP) and Elixhauser comorbidity score. A combination of the standard deviation of NN intervals (SDNN) as the HRV parameter, and FBT response in terms of the change in MAP (**Δ**MAP) as well as the change in SDNN (**Δ**SDNN) were then added hierarchically and also separately to Model-1 to develop additional models (Model-2 to Model-6). Finally, the performance of all the developed models were compared.

**Results:** A total of 5960 patients were screened and 542 were included following exclusion criteria. The model with a baseline traditional feature set (SAPS II, CPOF and **Δ**MAP) was surpassed by an augmented model (Model-6) using HRV measurements (SDNN and **Δ**SDNN) as additional features in terms of predictive performance (AUC: 0.871 vs 0.921, maximum F**_1_** score: 0.731 vs 0.792, indicating respective gains of 5.74% and 8.34%). In order of feature importance, following SAPS II score, **Δ**SDNN superseded **Δ**MAP.

**Conclusions:** The **Δ**SDNN can be an important predictor of mortality in septic patient requiring fluid bolus and is stronger than **Δ**MAP. Clinicians can choose **Δ**SDNN as an additional bedside parameter to predict mortality risk in sepsis patients.

## 1 Introduction

Sepsis, a life threatening organ dysfunction caused by dysregulated host response to infection, can quickly progress to septic shock and death, if left untreated [1, 2]. It is, therefore, imperative to analyze patient-specific efficacy of interventions such as fluid bolus therapy (FBT), a standard of care management for septic shock [3, 4]. At the same time, one seeks to accurately predict patient outcome so as to optimally allocate attention and resources to patients. Traditionally, severity score models including Simplified Acute Physiology Score (SAPS), Outcome Assessment Information Set (OASIS) and Sequential Organ Failure Assessment (SOFA) have found use in identifying high-risk patients. Patients present to the hospitals with baseline acute and chronic pathophysiological features, which, in their static values, form the basis of above models. In addition, one may argue that response patterns to basic and routinely used therapies can and should form part of prediction models, as a particular response to a therapeutic intervention simply reflects a patient’s underlying physiological reserve at the organ as well as the cellular level.

Although the justification of FBT in sepsis is not fully established [5], the probable rationale of the FBT is to raise venous volume relatively rapidly to stimulate the low-pressure volume receptors and also to preserve the intravascular volume, in the face of extensive capillary leakage. Fluid bolus now forms integral part of routine clinical care for patients with septic shock [6]. Therefore, FBT provides us with an unique opportunity to use it to examine a patient’s response to it at a deep physiological level and then to make that response an integral part of outcome prediction. It should be noted that none of the above mentioned models include therapy response as one of the predictive features [7–10].

It has been established that sepsis and septic shock can cause autonomic nervous system (ANS) dysfunction, resulting in impaired control of the heart and blood vessels and contributing to circulatory failure [11, 12]. There is dysfunction of ANS in septic shock due to desensitization and reduced responsiveness of high-pressure baroreceptors after prolonged stimulation, resulting in a maladaptive response to the hypotensive and inflammatory stress [13, 14]. Accordingly, cardiopulmonary baroreceptors, also known as volume receptors or atrial stretch receptors or low-pressure baroreceptors remains the available regulatory factor and a potential therapeutic target in septic shock.

Heart rate variability (HRV) analysis, which computes statistics on time intervals between successive cardiac beats, can be used to assess ANS activity [15, 16]. This has been previously shown to predict short and long term mortality in septic patients admitted to ICU [40, 41]. However, it is challenging to measure HRV in a clinical setting without manual intervention in real time. In this context, an algorithm has recently been suggested that enables automated accurate measurement of HRV by selecting a suitable window devoid of arrhythmia [17]. In the current study, we hypothesize that the prediction of mortality in sepsis patients can not only be improved by incorporating baseline HRV in the prediction model, but HRV response to FBT can also form a very strong predictive feature for hospital morality.

## 2 Methods

This retrospective cohort study adheres to the STrengthening the Reporting of OBservational studies in Epidemiology (STROBE) statement for reporting observational studies in epidemiology.

### 2.1 Data source and study population

Information, containing demographics, clinical parameters and outcomes, from sepsis patients treated in the ICUs of Beth Israel Deaconess Medical Center (BIDMC) between 2001 and 2012, were identified using ICD-9 codes (refer to Supplemental Table S1), from publicly available Medical Information Mart for Intensive Care III (MIMIC-III) & MIMIC-IV [18]. Further, the waveform matched subset was used to obtain ECG records linked to patients in the MIMIC clinical database [19, 20]. The use of MIMIC database was approved by the institutional review boards of both BIDMC and Massachusetts Institute of Technology (MIT), and informed consent was waived. Adult sepsis patients, who spent over 24 hours in the ICU and received an IV fluid bolus were included. This was defined by an infusion of predefined fluids (see Supplemental Table S2 for list of IV fluids) that were infused in less or equal to a 30-minute timeframe. We only included sepsis patients who underwent FBT within the first 48 hours of ICU admission. This timeline was chosen on the basis of previous work by Schmidt et al., [21]. We identified the earliest 5-minute window eligible for HRV analysis, as established in previously published work [17], one before (at time *t*_1_) and one after (at time *t*_2_) the administration of fluid bolus. The HRV parameter chosen for the study was computed corresponding to those windows (with respective values HRV*_t_*_1_ and HRV*_t_*_2_) and the change in HRV, ΔHRV=HRV*_t_*_2_ *−* HRV*_t_*_1_, was calculated. We excluded patients whose ECG records did not meet the eligibility criteria for HRV analysis [17]. As depicted by a flowchart in Figure 1(a), of the 5960 sepsis patients identified, 542 had matching waveforms.

**Fig. 1.**
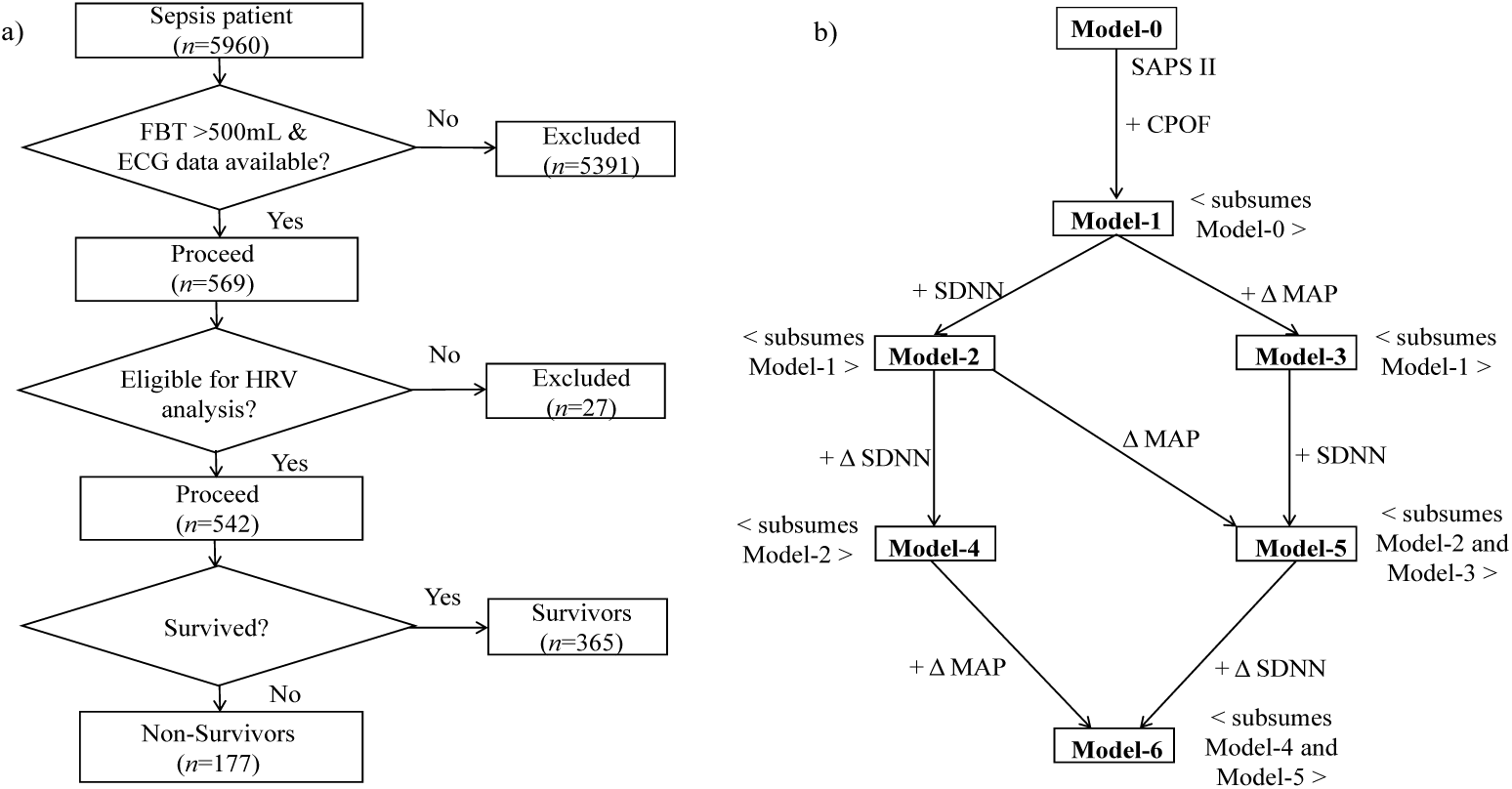
(a) Flow chart of selection of sepsis patients from MIMIC-III dataset. (b) Block diagram of proposed mortality prediction scheme. (CPOF: Common pool of features which includes elixhauser score, age, insurance, gender, ethnicity, admission type, HR, MAP; SDNN: Standard deviation of NN intervals (NNI); FBT: fluid bolus therapy; HR: heart rate; MAP: mean arterial pressure; ΔMAP: Change in MAP; ΔSDNN: Change in SDNN; *n*: number of sepsis patients. The baseline model (Model-0) was developed using SAPS-II as the foundational feature.)

### 2.2 Various predictors and models

The task of mortality prediction involves classification of a cohort into survivors and non-survivors, using available predictors/features. Based on clinical principles, data availability and findings from prior literature, we used the SAPS II score as the foundational predictor associated with mortality in sepsis patients [22, 23], and as the basis of our baseline model (Model-0). A common pool of features (CPOF), including patient demographics (age, sex, ethnicity), socioeconomic factors (insurance), admission type, chronic comorbidity (Elixhauser score), as well as heart rate (HR) and mean arterial pressure (MAP), were added to Model-0 to develop Model-1. Further advanced models incorporated features related to HRV and/or FBT response as explained subsequently.

HRV analysis, performed based on the RR intervals (RRI) of an electrocardiogram (ECG), should consider only normal RRI (NNI), because non-sinus beats are not subject to ANS activity in the same way sinus rhythm is [24]. We made use of the recently developed NAIHA algorithm to identify appropriate ECG windows and to compute SDNN, i.e., the SD of NNI^1^ [17]. The computed baseline (pre-FBT) SDNN at the eligible window at time *t*_1_, was added to Model-1 to develop Model-2.

In addition to change in MAP (ΔMAP) in response to FBT, [25] in this paper, we introduced an dynamic feature, namely, the change in SDNN, (ΔSDNN = SDNN*_t_*_2_ *−* SDNN*_t_*_1_), representing SDNN following and before an FBT respectively. Adding ΔMAP to Model-1, ΔSDNN to Model-2, and ΔMAP to Model-2, respectively, we would develop Model-3, Model-4 and Model-5. Finally, we would add ΔSDNN to the feature set of Model-5 to develop Model-6. Relationships among the various models under consideration are schematically depicted in Figure 1(b). Note that Model-0, Model-1 and Model-2 do not incorporate FBT response, while subsequent models do.

We used the NAIHA algorithm for HRV analysis [17], and extracted the remaining features directly using the pgAdmin PostgreSQL tool (v1.22.1) from the MIMIC-III clinical database.

### 2.3 Justification of the chosen HRV parameter

The analysis of consecutive difference of the SD of the means of RRIs involves examining how the SD values change from one segment to the next. This essentially reveals the rate or pattern of variability in the variability, so to say. Although SDNN is known to be influenced by multiple patient-based (e.g. supine position), therapeutic (e.g. drugs) and temporal (e.g. diurnal rhythm) factors [26–28], in the present context of measuring FBT response by ΔSDNN (= SDNN*_t_*_2_ *−* SDNN*_t_*_1_), the time interval *t*_2_ *−t*_1_ was approximately 30 minutes, a relatively short time, during which the aforementioned factors influencing SDNN can assumed be approximately constant. Further, we also performed similar analyses for representative frequency domain and as well as nonlinear domain [29], and the results have been reported.

### 2.4 Preparation of datasets

The dataset used in the study, consisting of 542 patient records, was randomly divided such that 434, i.e., 80% of the records formed the training set and the remaining 108, i.e., 20%, formed the test set. Using the training set (with 291 survivals and 143 non-survivals), a 5-fold stratified cross-validation were carried out to validate the ML models at hand. The test set remained unseen till this point. To prevent data leakage all data preprocessing steps were performed solely on the training set before validation.

### 2.5 AI architecture and performance indices

All predictive models at hand, namely, Model-0 and Models 1-6, were developed using a common AI architectural template. Each was based on an ensemble^2^ of successive decision trees (DTs) making use of eXtreme Gradient Boosting (XGBoost) [31]. We performed 5-fold stratified cross-validation on the training set to obtain the optimal combination of hyperparameters (see Supplemental Table S3). Such hyperparameters were chosen to maximize the average area under the (receiver operating characteristic, ROC) curve (AUC).

Subsequently, we compared the performance of the predictive models with AUC. Further, we performed decision curve analysis (DCA) [32, 33], where the variation in net benefits (clinical utility) was studied against a probability threshold (p_t_). We also assessed the real-world reliability of each model’s predictions in terms of calibration plots and calibration belts corresponding to the maximum F_1_ score [34]. We have also computed Sensitivity (Se), Specificity (Sp), accuracy, Net Reclassification Improvement (NRI) and Integrated Discrimination Improvement (IDI). Details regarding ROC, AUC, DCA, calibration plot, calibration belt, Se, Sp, NRI and IDI are provided in the Supplementary material. Finally, the features were ranked in the order of importance in making mortality prediction [35]. This was only done in the best performing model.

For each continuous features, we indicated the mean (SD), and tested the difference of its distributions within the rival (survivor/non-survivor) classes using the Kolmogorov-Smirnov test [36] (See Table 1). Categorical features were expressed as percentage, and the Chi-squared test was used to examine difference [37]. A *p*-value less than 0.05 was considered statistically significant. The SPSS software was used for statistical analyses. We used the winsorized method to handle outliers (extreme values). Specifically, we replaced extreme values below the 5th percentile and above the 95th percentile with the corresponding percentile values. Missing values constituted less than 2% of the dataset indicating a relatively small impact on our analysis. For numeric variables, we used mean imputation to replace missing values. For categorical variables, we used modal imputation, replacing missing values with the most frequently occurring category (mode).

**Table 1.**
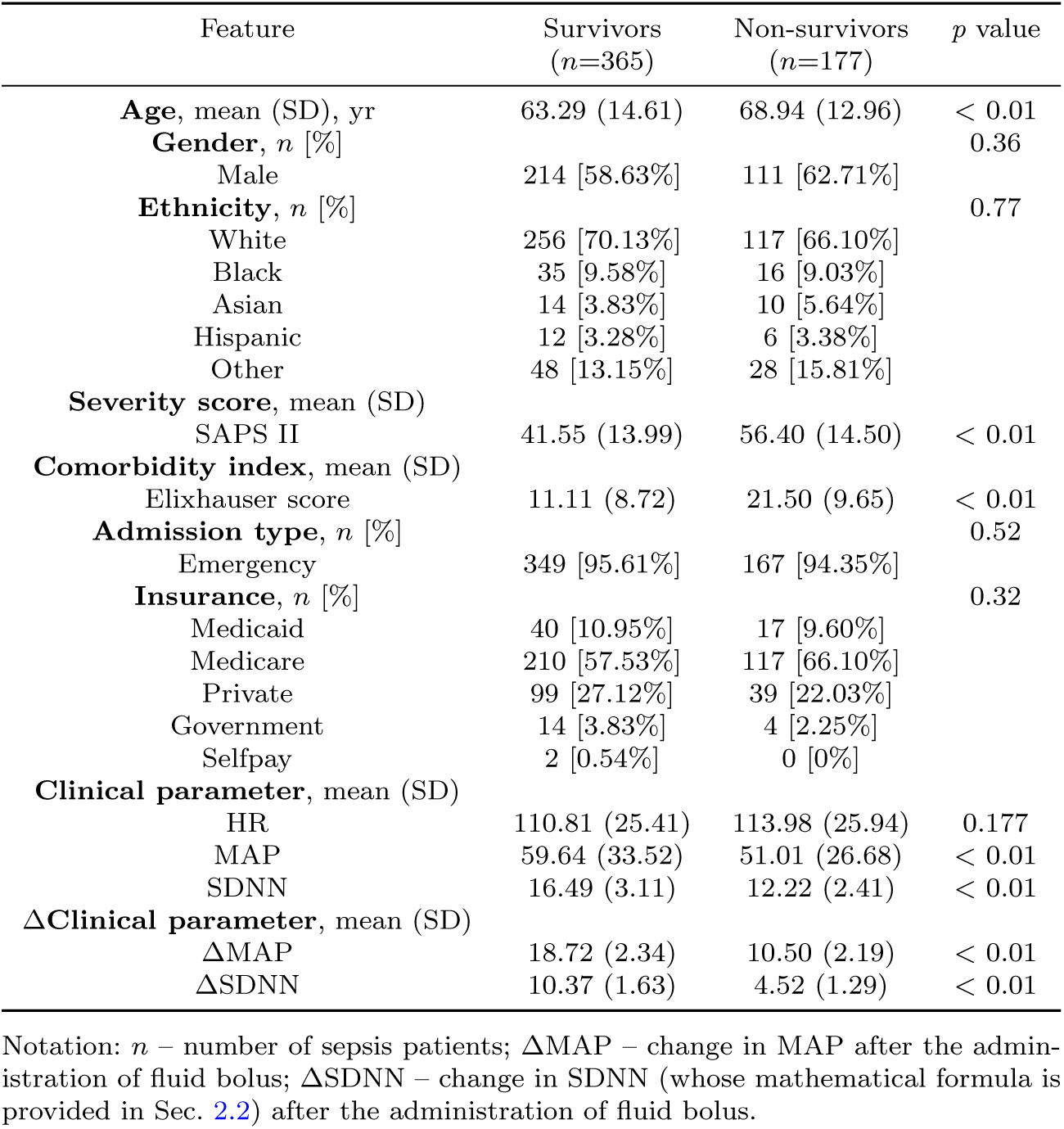
Patient statistics.

Codes were written in Python v3.8 and executed using resources from Google Colab. ML models were sourced from the scikit-learn v0.24 library, and optimized using Tensor Flow v2.

## 3 Results

Five hundred and forty two sepsis patient records with 365 survivors and 177 non-survivors were included in the analysis. Table 1 summarizes the patient demographics and acute and chronic clinical features. Significantly higher MAP (59.64 vs 51.01 mm Hg), SDNN (16.49 vs 12.22 ms), ΔMAP (18.72 vs 10.50 mm Hg), and ΔSDNN (10.37 vs 4.52 ms) values, as well as significantly lower SAPS II score (41.55 vs 56.40), Elixhauser score (11.11 vs 21.50), and age (63.29 vs 68.94) were seen in survivors. A positive value of ΔSDNN indicates an increased SDNN following FBT in both the groups of survivors and non-survivors. However, the increase was shown to be significantly more in survivor group (10.37 ± 1.63 vs. 4.52 ± 1.29; (*p <* 0.01)). The finding indicates that, while the FBT has got a general effect of increasing the rate of change in variability in the cardiac chronotropic activities, this action is markedly reduced in non-survivors.

The block diagram of proposed mortality prediction scheme is shown in Figure 1(b). The performance of baseline SAPS-II based Model-0 improved with the inclusion of CPOF in Model-1 (see Figure 2(a)). The inclusion of SDNN (Model-2) led to further improvement, but not as much as that of the model containing ΔMAP (Model-3). We next compared two separate models (Model-4 and Model-5), containing ΔSDNN and ΔMAP respectively with reference to Model-2. Addition of ΔSDNN (Model-4) caused larger improvement than the addition of ΔMAP (Model-5), indicating that the change in an HRV parameter is more informative as a predictor of mortality. Finally, model-6, containing the baseline acute and chronic patient based factors, along with ΔMAP, the baseline SDNN and ΔSDNN in response to FBT caused a considerable improvement in model performance, notably, 5.74% in AUC and 8.34% in maximum F_1_ score. We employed DeLong’s test to assess the statistical significance of differences in AUC between model pairs. The results yielded *p*-value less than 0.05 for all model pairs, indicating that each successive model iteration achieved a statistically significant improvement in predictive performance (refer to Supplemental Table S4).

**Fig. 2.**
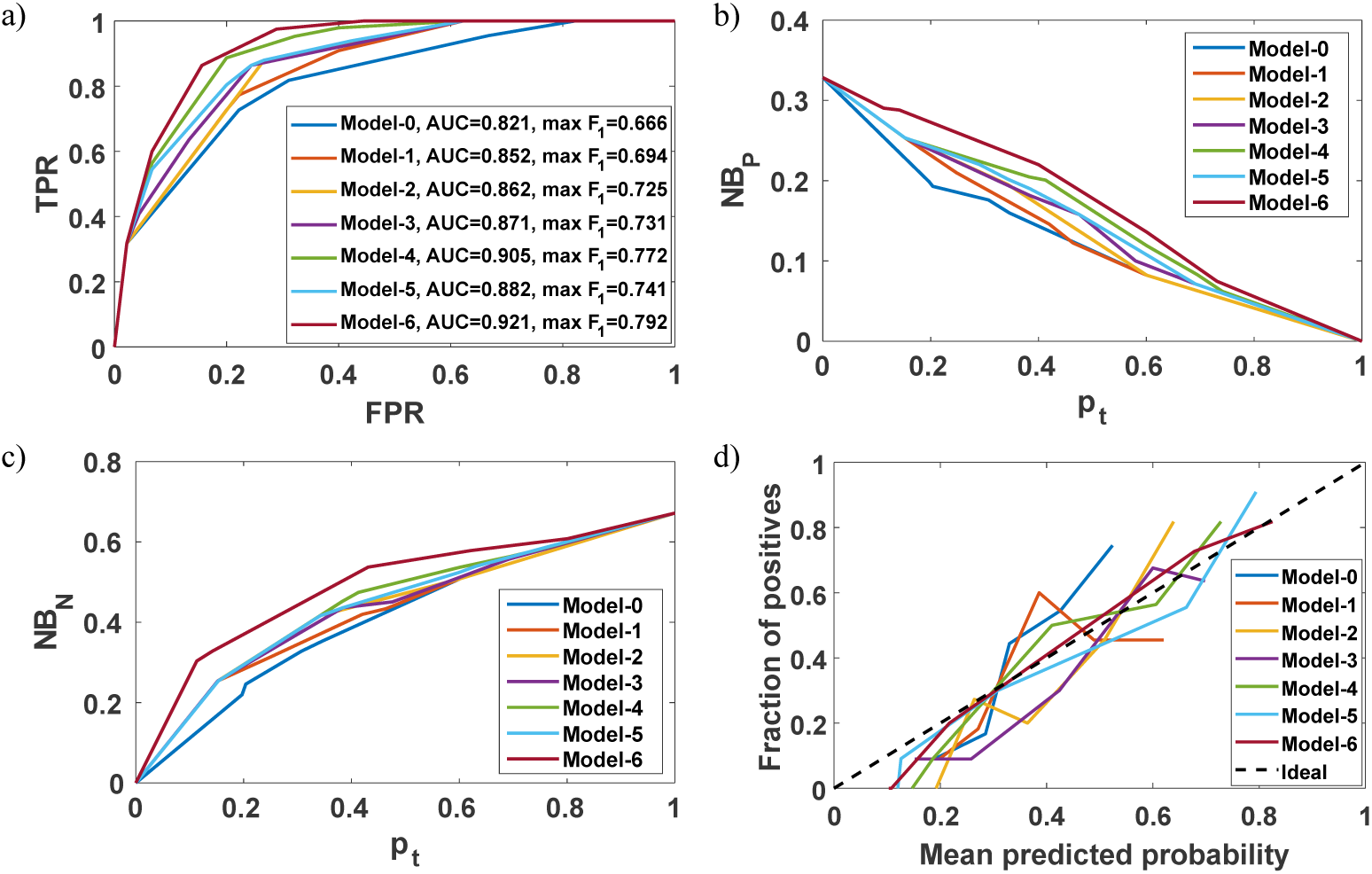
Performance of developed models with SDNN as HRV parameter. (a) ROC plots with operating point marked at max F_1_ score; Decision curve analysis for (b) positive/non-survived and (c) negative/survived predicted patients; (d) Calibration plot. (FPR: false positive rate, TPR: true positive rate, NB_P_: net benefit of positive predicted patients, NB_N_: net benefit of negative predicted patients, p_t_: probability threshold)

The additional performance metrics such as accuracy, Se, Sp, NRI and IDI are also provided (refer to Supplemental Table S5). A positive NRI values indicates that the new model correctly reclassifies a higher proportion of individuals compared to the baseline model, and a positive IDI values indicates that the new model has better discriminatory power than the baseline model.

Subsequently, a clinical utility based perspective is shown in Figures 2(b) and 2(c) in decision curve analyses. Model-6 outperforms other models by exhibiting higher net benefit for patients predicted both positive/non-surviving (NB_P_) and negative/surviving (NB_N_) across various probability thresholds (p_t_). Further, Model-6 (Figure 2(d)) exhibits satisfactory calibration, compared with other models. While the calibration-belt plots (Figure 3) demonstrated that each model somewhat underestimated the mortality risk of low-risk patients and overestimate the risk for high-risk patients, this was lesser in case of Model-6.

**Fig. 3.**
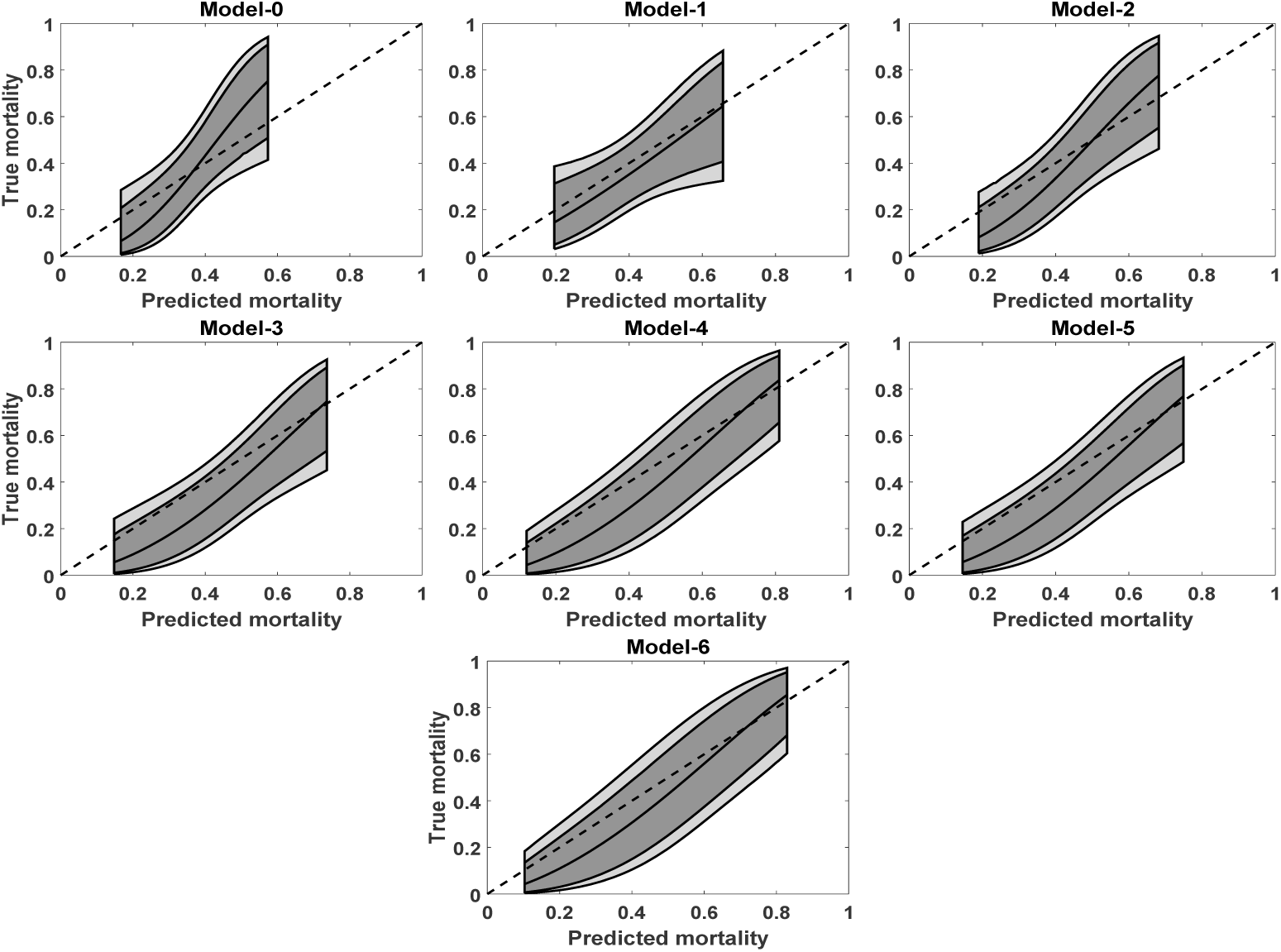
Calibration belts of developed models with SDNN as HRV parameter: Deviation from the bisector (45°dashed line of perfect calibration) at 80% (inner belt: dark area) and 95% (outer belt: light area) confidence levels.

Subsequently, we studied relative importance of various features in Model-6 as shown in Figure 4. SAPS II and Elixhauser scores, which represent large number of acute and chronic patient based factors respectively, unsurprisingly turned out to be most important. However, ΔSDNN proved to be a stronger feature compared to ΔMAP in the hierarchical importance. This finding is further supported by comparing the performance of Model-4 (includes ΔSDNN and not ΔMAP) and Model-5 (includes ΔMAP and not ΔSDNN), while keeping all other features the same in both the above models. Specifically, Model-4 achieved a higher performance (AUC: 0.905) compared to Model-5 (AUC: 0.882) indicating the superior predictive value of ΔSDNN than ΔMAP.

**Fig. 4.**
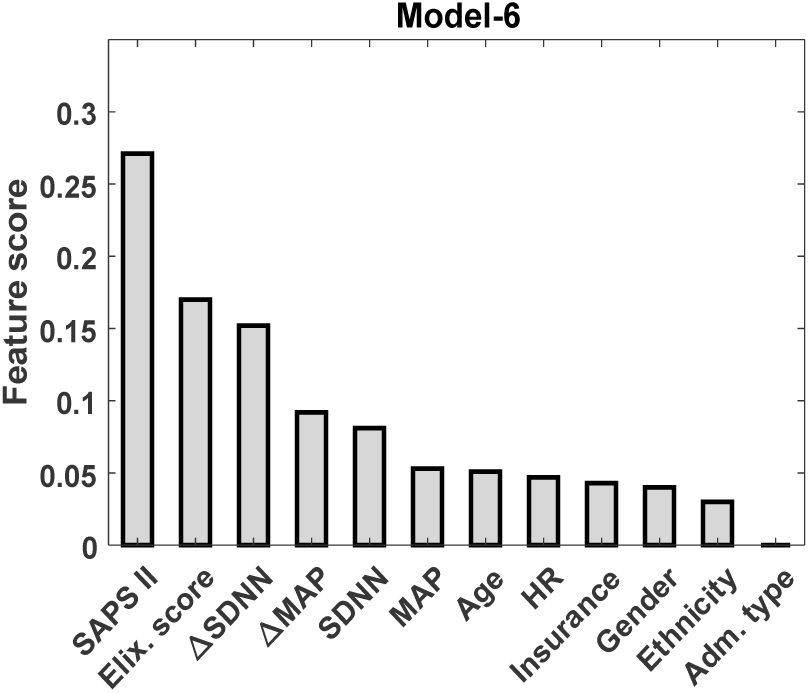
Feature importance plot of Model-6 with SDNN as HRV parameter.

## 4 Discussion

In this study, we developed AI models examining the effect of plasticity of HRV parameters on mortality following an intravenous fluid bolus in patients with sepsis. We incorporated established clinical scores and traditional bedside parameters in the model alongside baseline HRV and its change. While baseline HRV parameter has earlier been shown to be a predictor of mortality in sepsis, this is the first time it has been shown that higher ΔHRV, indicating better autonomic plasticity, could be a predictor for satisfactory outcome in this setting.

Furthermore, while we primarily validated our hypothesis that ΔSDNN can serve as a predictive feature for hospital mortality in sepsis patients based on the temporal HRV parameter SDNN, similar validation exercises can be conducted for HRV parameters in the frequency and nonlinear domains. Specifically, sets of Figures 5–7 and 8–10 depict comprehensive model performance analyses based, respectively, on frequency domain HRV parameter LF/HF (low frequency power to high frequency power) ratio^3^ and SD1/SD2 ratio^4^ The observed trends in each of the cases of LF/HF and SD1/SD2 ratios broadly align with those in the case of SDNN, generalizing our findings (additional performance metrics such as accuracy, sensitivity, specificity, NRI and IDI are provided in Supplemental Table S6 and S7).

**Fig. 5.**
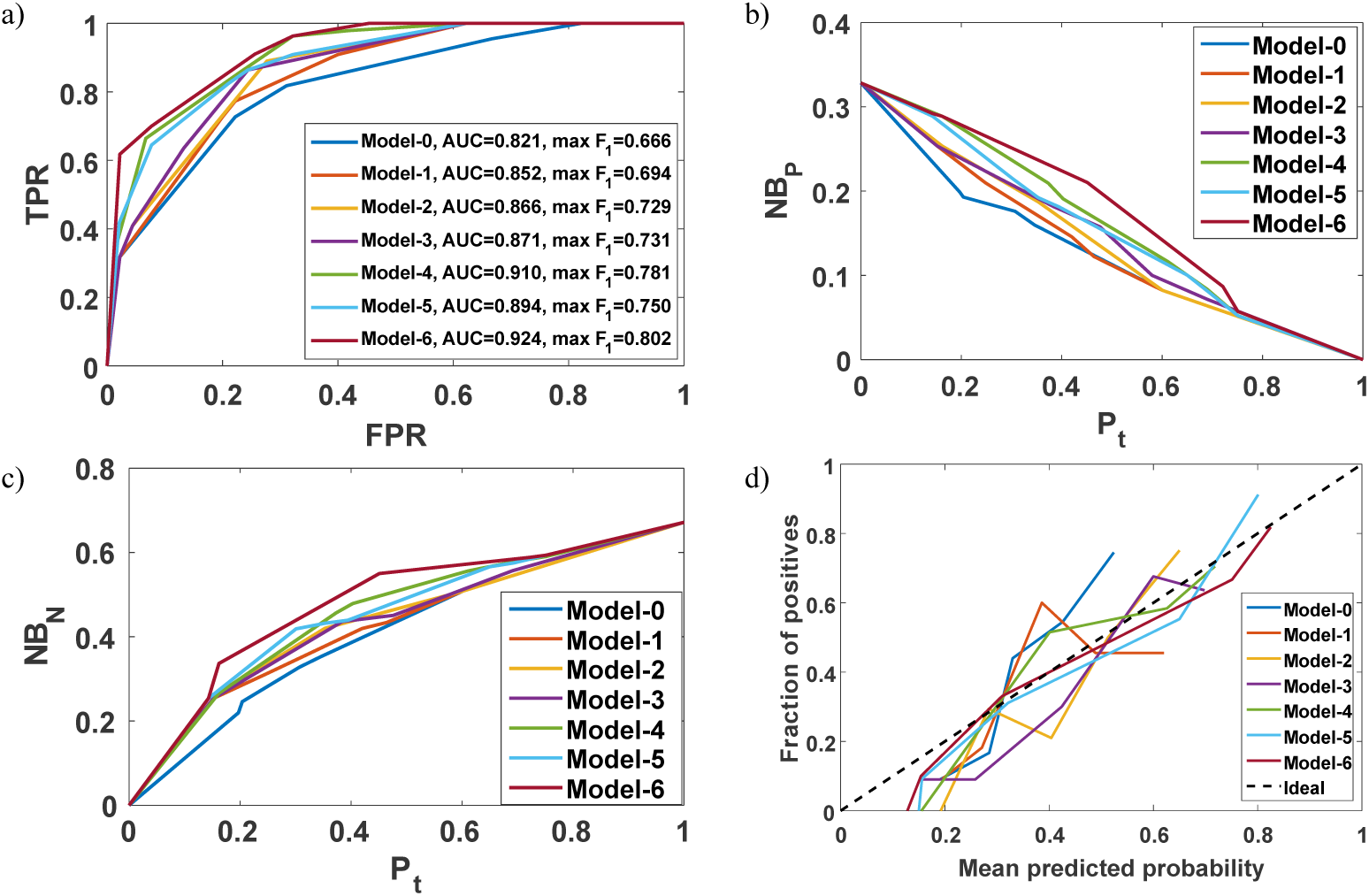
Performance of developed models with LF/HF ratio as HRV parameter. (a) ROC plots with operating point marked at max F_1_ score; Decision curve analysis for (b) positive/non-survived and (c) negative/survived predicted patients; (d) Calibration plot. (FPR: false positive rate, TPR: true positive rate, NB_P_: net benefit of positive predicted patients, NB_N_: net benefit of negative predicted patients, p_t_: probability threshold)

**Fig. 6.**
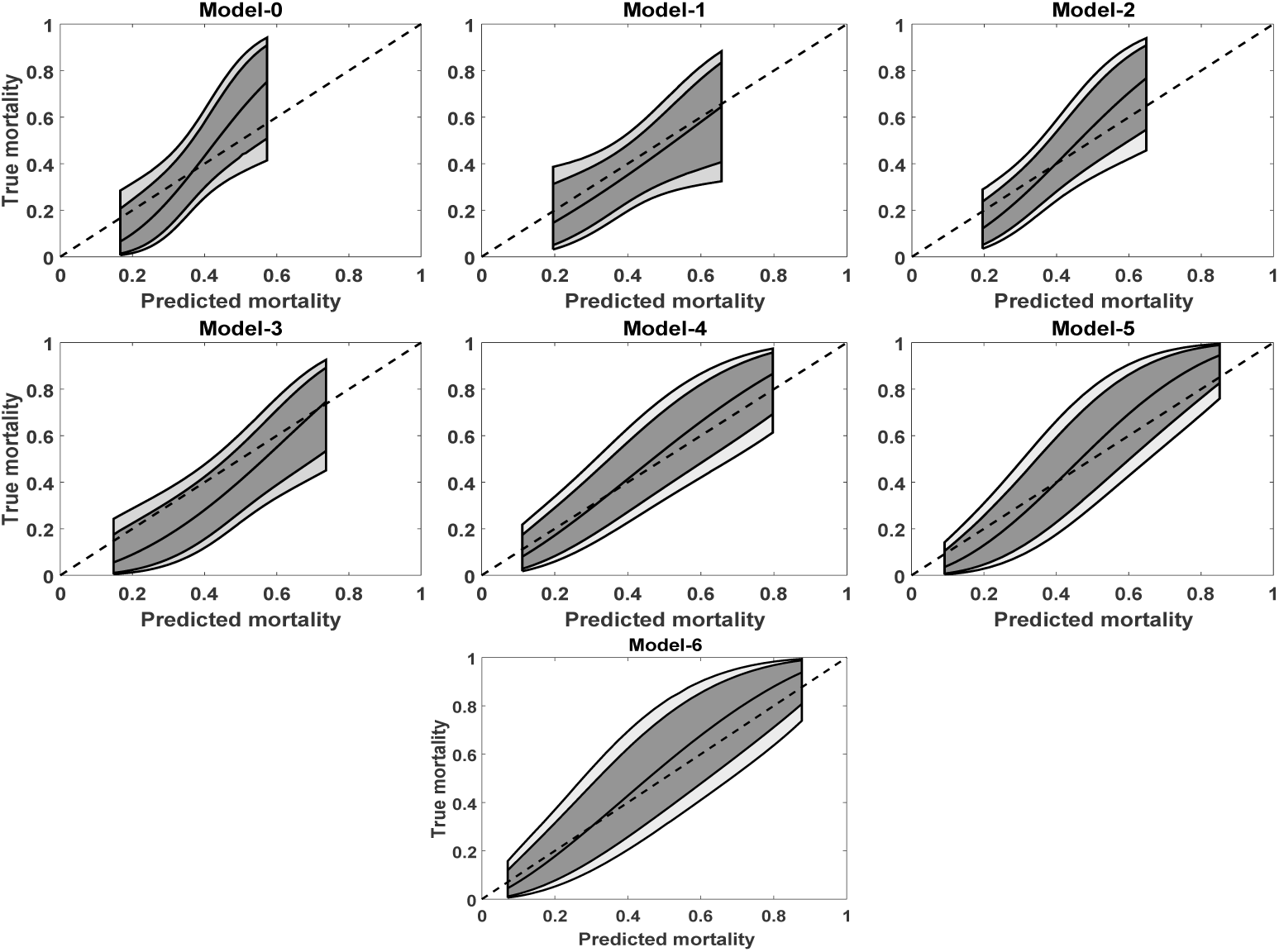
Calibration belts of developed models with LF/HF ratio as HRV parameter: Deviation from the bisector (45°dashed line of perfect calibration) at 80% (inner belt: dark area) and 95% (outer belt: light area) confidence levels.

**Fig. 7.**
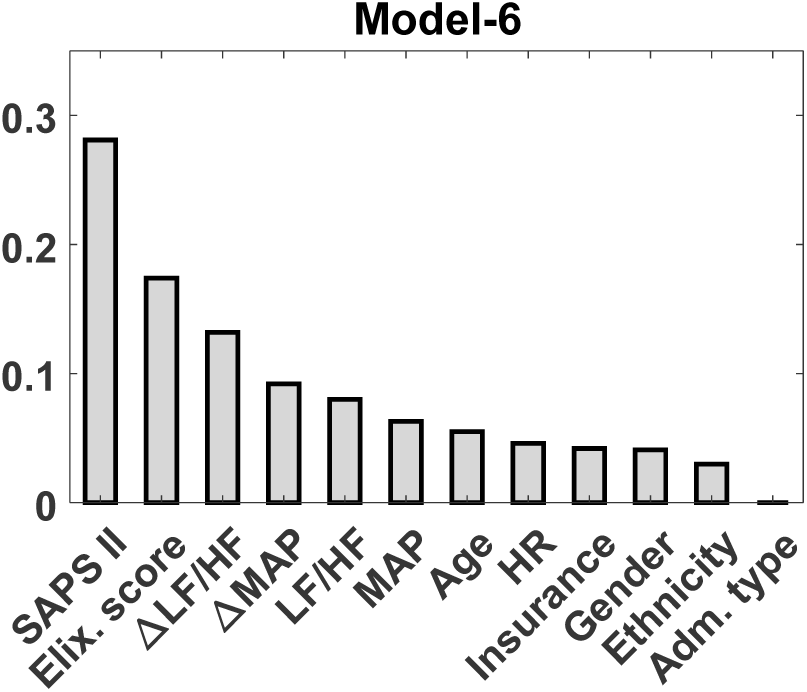
Feature importance plot of Model-6 with LF/HF ratio as HRV parameter.

**Fig. 8.**
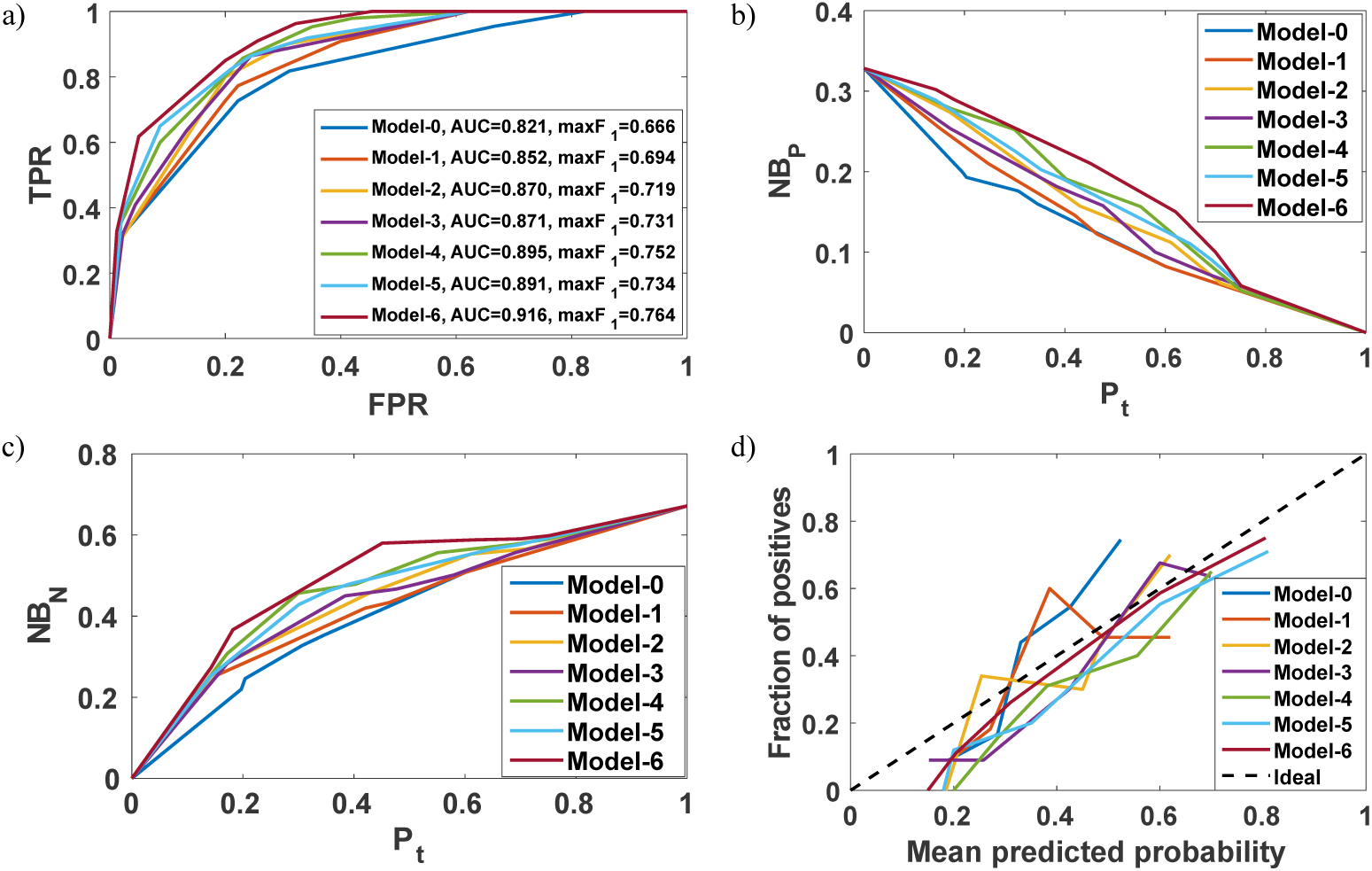
Performance of developed models with SD1/SD2 ratio as HRV parameter. (a) ROC plots with operating point marked at max F_1_ score; Decision curve analysis for (b) positive/non-survived and (c) negative/survived predicted patients; (d) Calibration plot. (FPR: false positive rate, TPR: true positive rate, NB_P_: net benefit of positive predicted patients, NB_N_: net benefit of negative predicted patients, p_t_: probability threshold)

We showed that ΔHRV is a stronger predictor of mortality than baseline HRV and other routinely used clinical parameters including ΔMAP (Figure 4). However, feature importance may vary among individual patients, which holds particular clinical significance. To gain insights into feature impact on individual patients, SHAP force plots were generated for different patients in Figure 11 [38]. SHAP force plots are a visualization tool that illustrates the impact of each feature on the predicted outcome for a particular instance, allowing us to identify the direction towards which each feature tends to drive the model’s predictions. By examining SHAP scores, one may gain insights into how (direction-wise) the model is using the input features to make predictions for individual patients. It should be noted that a positive SHAP score in this instance, given in bold, indicates a predicted status of non-survival, and negative SHAP score indicates a predicted status of survival in individual patient.

**Fig. 9.**
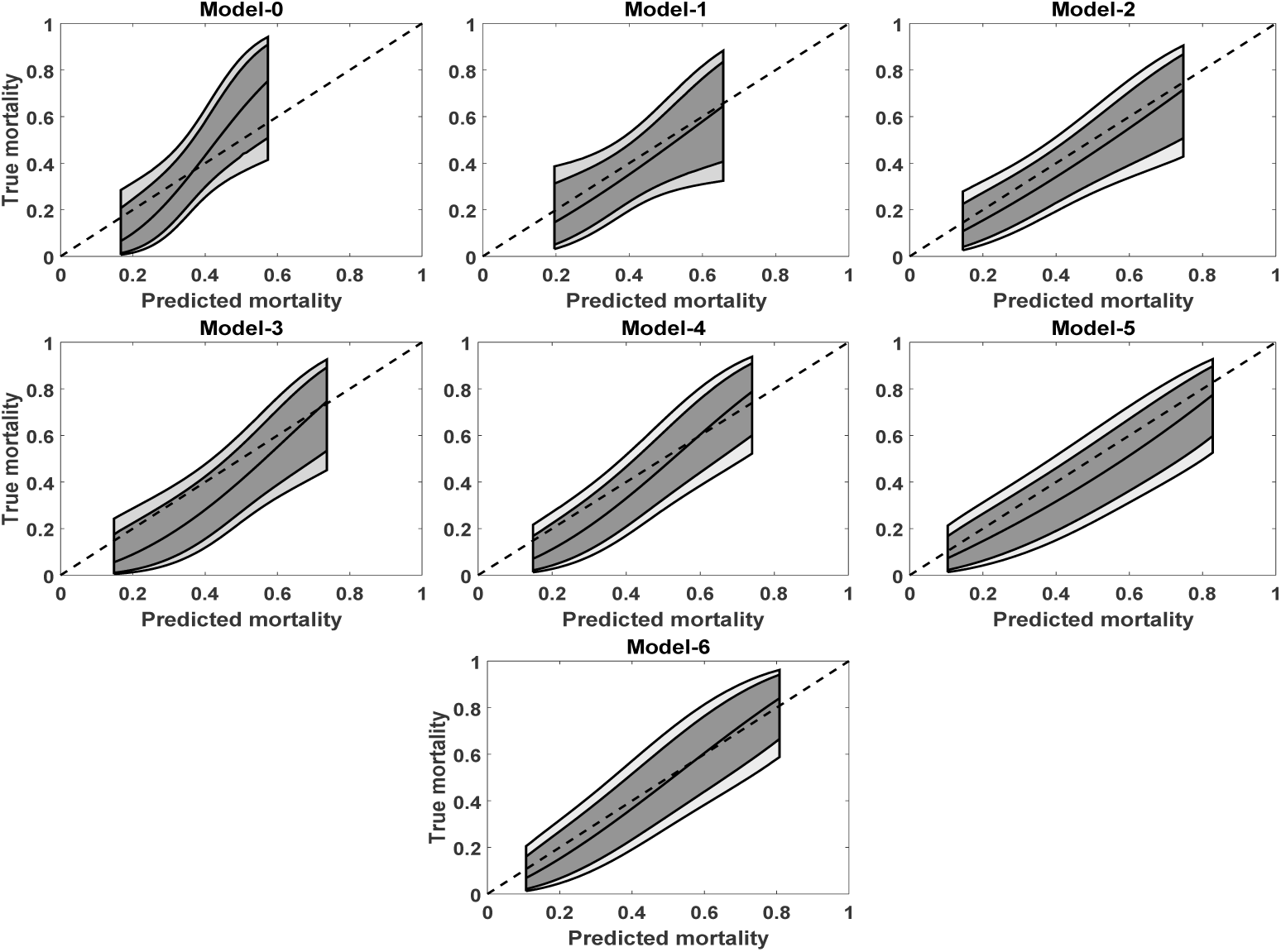
Calibration belts of developed models with SD1/SD2 ratio as HRV parameter: Deviation from the bisector (45°dashed line of perfect calibration) at 80% (inner belt: dark area) and 95% (outer belt: light area) confidence levels.

**Fig. 10.**
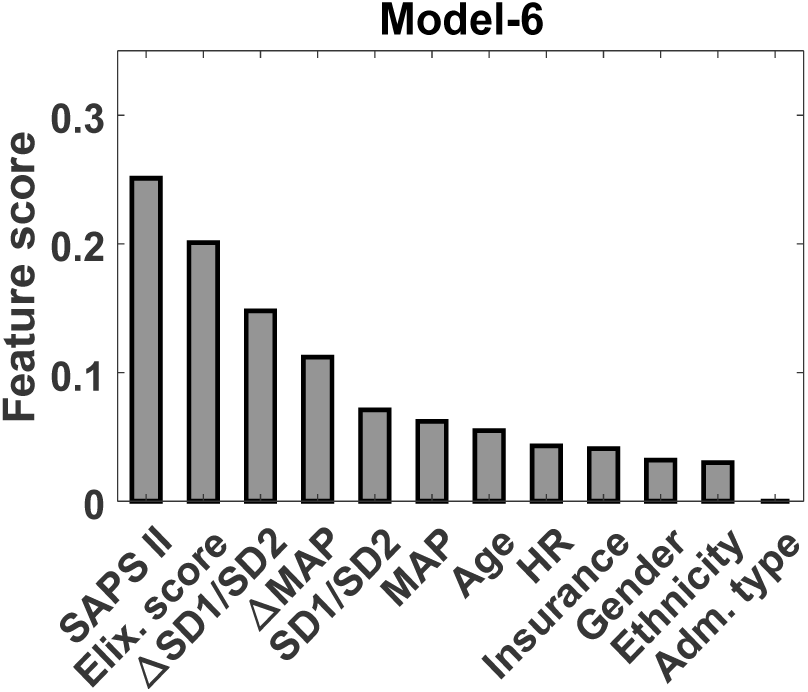
Feature importance plot of Model-6 with SD1/SD2 ratio as HRV parameter.

**Fig. 11.**
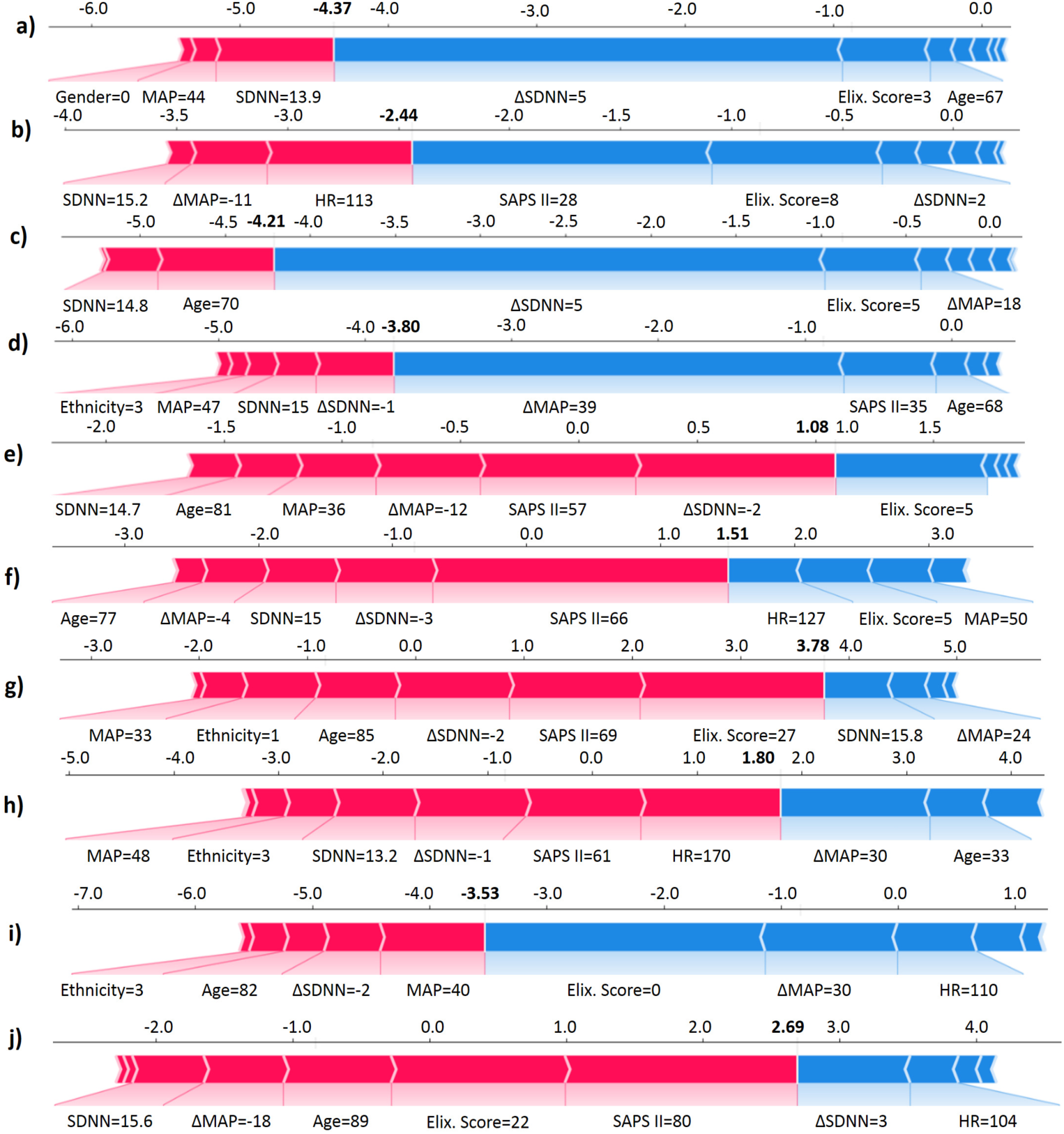
SHAP force plot of developed models for various cases from the test data. (a-d): survived patients predicted as survived (True negative); (e-h): non-survived patients predicted as non-survived (True positive); (i): non-survived patients predicted as survived (False negative); (j): survived patients predicted as non-survived (False positive).

A few representative cases of model prediction is shown in Figure 11. Figures 11(a-d) depict instances of correct predictions of survival, while Figures 11(e-h) showcase cases of correct predictions for death. There were a few instances where the model predicted the outcome incorrectly. In Figure 11(i), despite a ΔSDNN of -2, the low Elixhauser score (0) was strong enough to push the model to predict a status of survival. Similarly, in Figure 11(j), despite a ΔSDNN of 3, the high SAPS II score (80) leads the model to predict a status of non-survival. Therefore, ΔSDNN by itself, in these instances, proved to have the correct (direction-wise) influence on the model output, although other features dragged the overall prediction to the wrong direction. These local variations do not undermine the utility of ΔSDNN; instead, those highlight the nuanced and context-dependent nature of clinical prediction tasks.

Building on these observations, identifying a potentially useful threshold for ΔSDNN may support further investigation into its predictive value. Our analysis (see Figure 12) suggests that ΔSDNN thresholds of *≤* 4.5, *≤* 4.2, and *≤* 1.5 are associated with mortality risks of *≥* 10%, *≥* 20%, and *≥* 30%, respectively. The sensitivity/specificity metrics and the discussion on false positives/negatives are provided in the Supplement (refer Table S13). The choice of a particular threshold for ΔSDNN depends on various factors, such as patient age and comorbidities, resource availability, hospital settings, and clinical context. For example, in a limited resource condition, a clinician may choose a more conservative threshold. It is important to emphasize that the proposed ΔSDNN thresholds are exploratory and derived from the existing data. They were not intended for immediate clinical application or triage decisions, but rather to highlight patterns that warrant further investigations. We suggest these thresholds as preliminary reference points to guide future prospective studies aimed at validating their clinical relevance and generalizability.

**Fig. 12.**
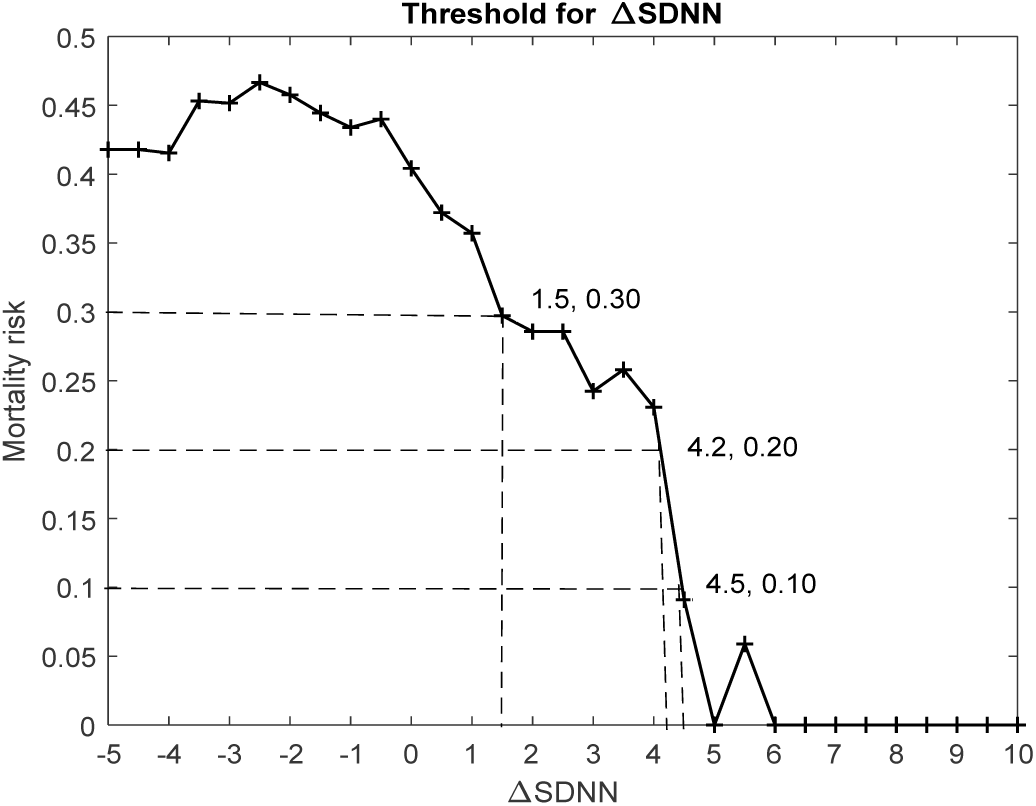
Mortality risk stratification based on ΔSDNN values. The choice of a particular threshold for ΔSDNN depends on various factors, such as patient age and comorbidities, resource availability, hospital settings, and clinical context.

It has been previously shown that impaired HRV in critically ill population is associated with increased mortality [21]. HRV represents the complex interaction between cardiac pacemaker cells and ANS. The loss of sympathetically and vagally mediated HRV is a risk factor for death in septic patients [41]. The possible explanation for this could be derived from the fact that vagal nerve stimulation, which improves HRV, attenuates multiple organ dysfunction in porcine sepsis model [42]. Overall reduction of HRV has been associated with pro-inflammatory state [43]. At the organic level, In a postmortem study, greater degrees of ischemia and neuronal and glial apoptosis were observed in cardiovascular autonomic centers of patients with septic shock than those without septic shock [44]. At the molecular level, there is evidence of upregulation of G-protein–coupled receptor kinase 2 expression in cardiomyocytes in autonomic dysfunction, leading to impaired cardiac function [45]. It should also be noted that HRV can be diminished by endotoxemia prevalent in sepsis [47, 48]. Baroreceptor dysfunction, which is closely associated with autonomic function and HRV, has previously been shown to be associated with poor outcome in patients with multiorgan failure when dysfunctional [49]. Additionally, in patients receiving goal directed therapy (GDT), signs of tissue hypoxia was worse if they had baroreceptor dysfunction [50]. It has been explained that, persistently high sympathetic drive may lead to saturation of low-frequency oscillatory systems [51], or that excessive concentrations of circulating catecholamines may compromise central autonomic control [52, 53]; similar to the situation in patients with heart failure. Therefore there is adequate evidence of interplay between ANS and inflammation, that can influence patient outcome.

Surviving sepsis guideline advocates sepsis-6, which includes fluid bolus as one of the major components [6]. Our study shows that fluid bolus can also be used, in addition to being a resuscitative intervention, to identify what can be termed as autonomic reserve in septic patients in the form of ΔHRV and thereafter in risk stratification. This can potentially be used not only for patients in the ICU but also for patients in the emergency departments in order to triage the sicker ones to critical care or enhanced care units instead of transferring them to the ward. This needs to be tested in prospective septic patient cohorts in the future. It is important to emphasize that the findings of this study do not attribute causality of mortality on HRV plasticity, but highlights an association of reduced autonomic reserve with poor clinical outcome, and how that can be used to stratify patients.

We would like to emphasize that we do not want to propose ΔHRV as a marker for fluid responsiveness. We are not proposing a certain ΔHRV as a treatment target either, but purely as a marker for poor underlying physiology. The usual set of parameters that are used at the bedside in the real world to risk-stratify shocked patients are serum lactate and capillary refill time. The dynamics of both of these parameters have been associated with poor patient outcome. Blood pressure is also used as a target outcome for fluid bolus. Given the fact the fluid bolus is used in literally every patient with septic shock, we used this as a physiological stress test to examine the HRV plasticity, which in turn reflects autonomic plasticity.

There are certain limitations in our study. First, serum lactate, which is a strong predictor of mortality in sepsis, could not be included due to unavailability of this parameter consistently around the time of FBT. Second, the current study assumes that all other relevant factors/confounders (such as sedation, ventilation status, vasopressor use, intrinsic physiological rhythms, and drug effects) affecting HRV remain relatively stable before and after the short time span of FBT and therefore their effect on HRV parameters remains the same in those time points. Although, we leveraged the MIMIC waveform database’s continuous recordings and accurately computed HRV changes using clinician-logged time points for fluid bolus administration, potential small uncertainties in these time points may exist. Given that the selection of the patients within the waveform database from the wider MIMIC is not based on a particular declared selection criteria, we assume that their is no particular selection bias. However, it is possible that there were unknown factors that drove the patient selection within the waveform dataset. If this was the case, the generalizability of our findings would accordingly be limited.

In future, a targeted study needs to be specifically designed to identify how different sepsis subphenotypes respond to FBT, considering the potential for distinct relationships between ΔHRV and outcome across subphenotypes. This could involve applying methods to identify subphenotypes [54], and determining the most suitable HRV parameter for evaluating FBT response in each subgroup. Further research needs to be done to determine the range of change of HRV that holds clinical significance. Future work should also involve developing interpretable models that go beyond the current feature set (see Figure 4), including lactate dynamics and interplay of different medications. Finally, various patient subgroups (such as diabetes, cardiac failure etc.) need to be examined to identify which ones, if any, benefit significantly from this approach. Most importantly, this may open the door for actionable AI to be used at the bedside for the critical care physician by making this real-time deeper physiological parameter available for clinical decision making.

## 5 Conclusions

In this retrospective study, improvement in sepsis mortality prediction was achieved by incorporating ΔHRV as an additional bedside parameter to assess ANS activity and evaluate the response to FBT. While our findings suggest that HRV dynamics can offer added predictive value, further exploration of this should be conducted via multicenter prospective studies before utilizing this as a clinical triage tool.

## Data Availability

All data produced are available online at https://mimic.physionet.org/

## Abbreviations

AI: Artificial Intelligence
AUC: Area under the ROC curve
ROC: Receiver operating characteristics
BIDMC: Beth Israel Deaconess Medical Center
MIMIC: Medical Information Mart for Intensive Care
MIT: Massachusetts Institute of Technology
CPOF: Common pool of features
MAP: Mean arterial pressure
RRI: R-R interval
ECG: Electrocardiogram
NAIHA: Novel AI based HRV Analysis
SDNN: Standard deviation of N-N interval
DT: Decision tree
XGBoost: eXtreme Gradient Boosting
DCA: Decision curve analysis
SD: Standard deviation
NB: Net Benefit
FBT: Fluid bolus therapy
HRV: Heart rate variability
SAPS: Simpli-fied Acute Physiology Score
SOFA: Sequential Organ Failure Assessment
OASIS: Outcome Assessment Information Set
ANS: Autonomic nervous system
STROBE: STrengthening the Reporting of OBservational studies in Epidemiology.

## Declarations

### Ethics approval and consent to participate

Consent obtained for use of MIMIC databases.

### Consent for publication

Not applicable.

### Availability of data and materials

Publicly available datasets were used in this study. The MIMIC dataset is available at https://mimic.physionet.org/.

### Authors’ contributions

R.L.R. developed the algorithms, performed the experiments, prepared an initial draft and revised it several times based on inputs. R.S. and A.S. provided clinical inputs, clinical interpretation and critical review. S.J. and R.S. provided overall supervision, approved study design, algorithms and results, and finalized the manuscript.

## Acknowledgements

Not applicable.

## Footnotes

1 Given a time series of *n* NNIs {*NN*_1_, *NN_2_*, …*NN_n_*}, one computes the standard deviation as 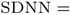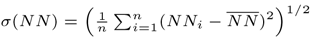, where 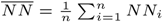 denotes the mean and *σ*(*NN*) denotes the standard deviation of the time series *NN*.

2 In general, ensemble of DTs significantly outperform single DT [30].

3 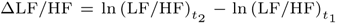, where 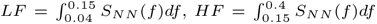, and *S_NN_* (*f*) denotes the power spectral density of time series *NN*.

4 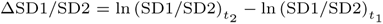, where *SD*1 = *σ*(*d*_1_), *SD*2 = *σ*(*d*_2_), 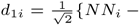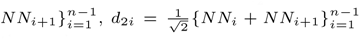 and *σ*(*·*) indicates the standard deviation of the argument time series.

